# COHD-COVID: Columbia Open Health Data for COVID-19 Research

**DOI:** 10.1101/2020.11.17.20232983

**Authors:** Junghwan Lee, Jae Hyun Kim, Cong Liu, George Hripcsak, Casey Ta, Chunhua Weng

## Abstract

Massive research efforts have been made in response to the COVID-19 (coronavirus disease-2019) pandemic. Utilization of clinical data can accelerate these research efforts to fight against the pandemic since important characteristics of the patients are often found by examining the clinical data. To provide shareable clinical data to catalyze COVID-19 research, we present Columbia Open Health Data for COVID-19 Research (COHD-COVID), a publicly accessible database providing clinical concept prevalence, clinical concept co-occurrence, and clinical symptom prevalence for hospitalized COVID-19 patients. COHD-COVID also provides data on hospitalized influenza patients and general hospitalized patients as comparator cohorts. The data used in COHD-COVID were obtained from Columbia University Irving Medical Center’s electronic health records. We expect COHD-COVID will provide researchers and clinicians quantitative measures of COVID-19 related clinical features to better understand and fight against the pandemic.

## BACKGROUND & SUMMARY

The novel coronavirus disease-2019 (COVID-19) has threatened the health of tens of millions of people all over the world. The global pandemic caused by COVID-19 has sparked massive research efforts in the fight against the novel disease, including characterizing the disease and clinical progression, identifying risk factors for hospitalization, and finding drugs that can be repurposed to lessen disease severity^1-3^. Utilization of clinical data from different institutions, hospitals, and nations can accelerate these research efforts since important characteristics of the patients are often found by examining the shared clinical data. Although many studies sharing epidemiological data^4,5^, public health^6^, and social measures^7^ for COVID-19 research have been conducted, publicly accessible clinical data on COVID-19 remain limited despite the immediate need^8^.

Columbia Open Health Data (COHD) provides open access to prevalence and co-occurrence statistics on conditions, drugs, procedures, and demographics derived from structured electronic health records (EHR) from Columbia University Irving Medical Center (CUIMC)^9^, which serves the large and diverse population of New York City (NYC) and its surrounding areas. Since its deployment, COHD has accelerated biomedical research by providing two informative resources, prevalence and co-occurrence statistics, and their derived association metrics^9-11^.

New York City has been one of the first epicenters of the disease in the United States since the first COVID-19 patient was confirmed on February 29, 2020^12^. As one of the largest hospitals in NYC, CUIMC has admitted more than 4,000 patients as of September 1, 2020. With the aim to provide shareable clinical data to catalyze future COVID-19 research, we extend our efforts on COHD and present Columbia Open Health Data for COVID-19 Research (COHD-COVID), a publicly accessible database providing clinical concept prevalence, clinical concept prevalence ratio between cohorts, clinical concept co-occurrence, and clinical symptom prevalence for COVID-19 patient cohort and comparator cohorts (hospitalized influenza patient cohort and general hospitalized patient cohort) derived from CUIMC’s electronic health records. In addition to providing accessible data, we also developed COHD-COVID web application programming interface (API) for easy access and better usability (available at http://covid.cohd.io).

## METHODS

We use the term “concept” to refer to clinical entities and events such as conditions (i.e., diagnosis), drugs, and procedures. The concepts and their names are defined by the Observational Medical Outcomes Partnership (OMOP) Common Data Model (CDM). When concepts are referenced in this paper, the name of the concept is styled in italics (e.g., *Disorder of respiratory system*) to distinguish the formalized concepts from regular text. We also styled entities in the OMOP CDM (e.g., *person_id* column in *condition_occurrence* table) in italics.

**Figure 1** depicts the overall workflow to create COHD-COVID. Columbia’s clinical data warehouse has been converted to the OMOP CDM on a regular basis. We first filtered the EHR data according to the cohort definitions, then EHR for each patient’s inpatient visits were identified. Condition, drug, and procedure concepts were obtained from the identified visits. Concept prevalence, concept prevalence ratio, concept co-occurrence, and symptom prevalence analyses were followed using the obtained concepts. To protect patient privacy, we excluded rare concepts observed in 10 or fewer visits and perturbed the true counts using Poisson randomization. Perturbed counts generated by the Poisson randomization process do not show statistically significant difference from the true counts^9^. The resulting data were stored in a MySQL database and made publicly available via a web API (http://covid.cohd.io/).

**Figure 1.**
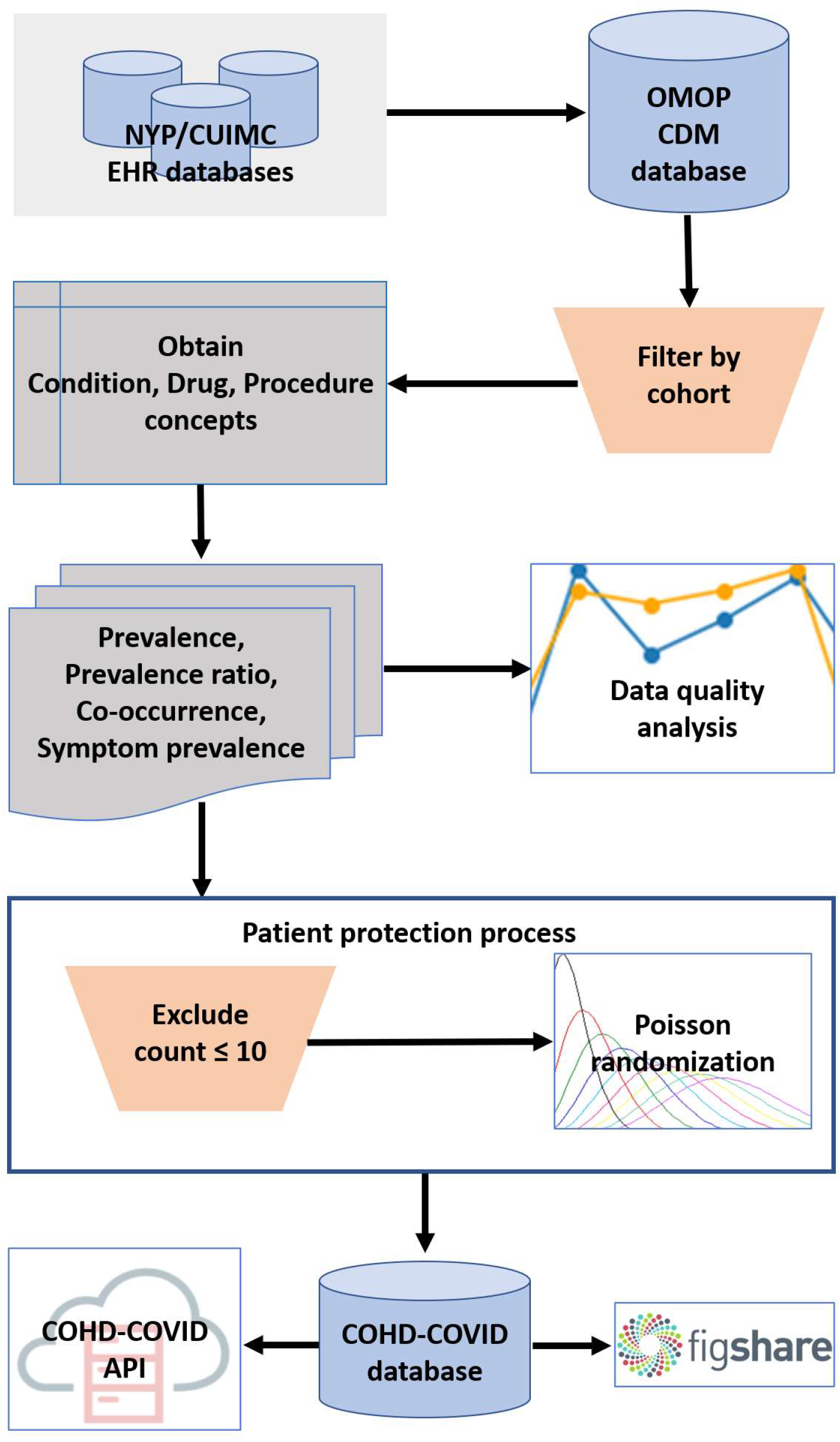
Overall workflow of Columbia Open Health Data for COVID-19 Research (COHD-COVID)

### Data Source

We utilized EHR data from the CUIMC’s clinical data warehouse, where inpatient and outpatient data dating back to 1985 are stored. CUIMC has converted its clinical data warehouse to OMOP CDM on a regular basis. Since CUIMC covers New York City and the surrounding area, which has a diverse population of 8.2M people and was an early COVID-19 epicenter, the EHR data from CUIMC can provide a diverse and large sample of COVID-19 patients.

Three different patient cohorts were used in this study. The COVID-19 cohort was defined as hospitalized patients aged 18 or older with a COVID-19 related condition diagnosis and/or a confirmed positive COVID-19 test during their hospitalization period or within the prior 21 days. Patients identified with the COVID-19 cohort definition from March 1, 2020 to September 1, 2020 were used for the COVID-19 cohort. The influenza cohort was similarly defined as patients aged 18 or older who had at least one occurrence of influenza conditions or pre-coordinated positive measurements or positive influenza testing in the prior 21 days or during their hospitalization period. The general cohort was defined as all hospitalized patients aged 18 or older. Patient visits from calendar years 2014 to 2019 were included for the influenza and general cohorts. The COVID-19 cohort was divided into sub-cohorts stratified by sex (male vs. female) and age (adult (18-64) vs. senior (65+)) for further investigation. The COVID-19 and influenza cohort definitions were adapted from the cohort definitions created by the Observational Health Data Science and Informatics’ (OHDSI) international network study for COVID-19^3^.

Patients belonging to the cohorts based on the three cohort definitions above were identified using the unique *person_id* from the *person* table in the OMOP CDM. Condition, drug, and procedure concepts observed in these patients during inpatient visits were extracted from the *condition_occurrence, drug_exposure*, and *procedure_occurrence* tables in the OMOP CDM, respectively. Inpatient visits of patients and the concepts in these visits were identified using *visit_occurrence_id* and *person_id*, along with the *visit_concept_id* from the *visit_occurrence* table. We used visit-based count instead of patient-based count in the following analyses for robust comparison between cohorts lie in different observation window. For example, if we use patient-based count, the patients included in cohorts with longer observation window are likely to have more concepts than the patients in cohorts with shorter observation window, which could inject bias into the metrics used in the analyses. Thus, we used visit-based count that cannot be affected by the length of observation window to reduce the bias.

### Concept Prevalence and Concept Prevalence Ratio Analysis

We calculated the concept prevalence in each cohort as detected from the EHR. The concept prevalence is defined as Eq(1):

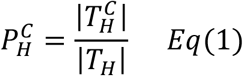

where 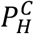 is the prevalence of concept *C* in cohort 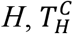 is the set of unique inpatient visits of patients in cohort *H* observed with concept *C*, and *T*_*H*_ is the set of unique inpatient visits of patients in cohort *H*. We also calculated hierarchical concept prevalence by defining 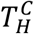 as the set of unique visits of patients observed with concept *C* or any of concept *C*’s descendant concepts as defined in the *concept_ancestor* table in the OMOP CDM. For example, the hierarchical count for *Ibuprofen* (OMOP concept ID 1177480) not only includes entries where the specific concept *Ibuprofen* was used, but also includes entries using other descendant concepts, such as *Ibuprofen 600 MG Oral Tablet* (OMOP concept ID 19019073). Taking hierarchical relationships into account mitigates some of the issues with coding variations across time and practices as different concepts with minor semantic differences can be aggregated into higher-level concepts.

The concept prevalence ratio indicates how frequently concept *C* occurs in cohort *A* relative to cohort *B*. The natural logarithm of the concept prevalence ratio is defined as Eq(2):

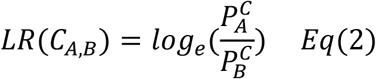

where *LR(C*_*A,B*_) is the log ratio of the prevalence of concept *C* for cohort *A* to cohort 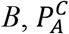 is prevalence of concept *C* in cohort *A*, and 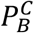 is prevalence of concept *C* in cohort *B*. Hierarchical concept prevalence ratio can be calculated by using hierarchical concept prevalence 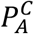 and 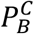.

### Concept Co-occurrence Analysis

Concept co-occurrence represents how frequently a specific concept pair appears in a cohort. We defined concept co-occurrence prevalence as Eq(3):

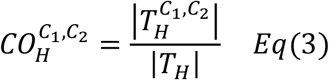

where 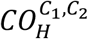 is the co-occurrence prevalence of concepts *C*_1_ and *C*_2_ in cohort 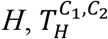 is the set of unique visits of patients observed with concept *C*_1_ and *C*_2_ in cohort *H*, and *T*_*H*_ is the set of unique visits of patients in the cohort *H*. We also calculated hierarchical concept co-occurrence using the hierarchy of concepts as described above.

### COVID-19 Symptom Prevalence Analysis

Since clinical symptoms often include multiple granular clinical concepts, a set of related concepts for a symptom is required to calculate the prevalence of the symptom. For example, dyspnea, which is one of the major symptoms of COVID-19, can be detected as standard concept *Dyspnea* or *Acute respiratory distress* in different patients. The two concepts do not have any hierarchical relationship and thus will not be aggregated by the hierarchical prevalence analyses. Thus, a concept set containing both *Dyspnea* and *Acute respiratory distress* is needed for accurate calculation of the prevalence of cough symptom. We defined symptom prevalence as Eq(4):

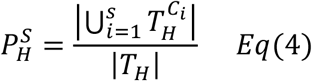

where 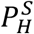 is prevalence of symptom *S* in cohort 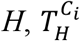 is the set of unique visits of patients observed with concept *C*_*i*_, *T*_*H*_ is the set of unique visits of patients observed in the cohort *H*, and *s* is the number of the unique concepts in the concept set for symptom *S*. Hierarchy between concepts is not considered in the symptom prevalence analysis since a concept set of a symptom already reflects hierarchy for that symptom.

We used concept sets for 11 major symptoms of COVID-19 (Cough, Chills, Abdominal pain, Diarrhea, Dyspnea, Fatigue, Fever, Myalgia, Nausea and vomiting, Tachypnea, and Throat pain) that have been defined by OHDSI to calculate symptom prevalence. The concepts included in each symptom are available in figshare^13^.

### Data Records

The results of all analyses, concept definitions, and concepts included in each symptom are available as tab-delimited flat files in figshare^13^. The results of the concept prevalence ratio analysis were not included in the record since they can be directly computed from concept prevalence available in the record. The COHD-COVID API (http://covid.cohd.io/) provides the results of concept prevalence and concept co-occurrence analyses. The prevalence and co-occurrence are relative to a maximum of 1.0 (1.0 = 100%) and all concepts are referenced by their OMOP standard concept ID. Brief descriptions for the flat files are:

#### covid_full_(non)hierarchical_concept_prevalence.txt

Hierarchical and non-hierarchical concept prevalence for the full COVID-19 cohort. The columns are OMOP concept ID, count of patients’ visits with the concept, and prevalence of patients’ visits with the concept. Concept prevalence for all sub-cohorts of the COVID-19 cohort are also available using the same file structure and with the sub-cohort name indicated in the filename (i.e., covid_[adult|senior|male|female]_(non)hierarchical_concept_prevalence.txt.)

#### covid_full_(non)hierarchical_concept_cooccurrence.txt

Hierarchical and non-hierarchical concept co-occurrence prevalence for the full COVID-19 cohort. The columns are the first OMOP concept ID, second OMOP concept ID, count of patients’ visits with the concept pair, and prevalence of patients’ visits with the pair of concepts. Concept co-occurrence prevalence for all sub-cohorts of the COVID-19 cohort are also available using the same file structure and with the sub-cohort name indicated in the filename (i.e., covid_[adult|senior|male|female]_(non)hierarchical _concept_cooccurrence.txt).

#### covid_full_symptom_prevalence.txt

Symptom prevalence for the full COVID-19 cohort. The columns are symptom name, the number of unique concepts having nonzero count in the symptom, count of patients’ visits with the symptom, and prevalence of patients’ visits with the symptom. Symptom prevalence for all sub-cohorts of the COVID-19 cohort are also available (i.e., covid_[adult|senior|male|female]_symptom_prevalence.txt).

#### covid_epic_shift_pvalue.txt

p-values of t-tests between counts from pre-Epic switch period and post-Epic switch period for all concepts in the full COVID-19 cohort. The columns are OMOP concept ID and p-value of the t-test for the concept. These are results from part of the data quality analysis in the following section.

#### influenza_(non)hierarchical_concept_prevalence.txt

Hierarchical and non-hierarchical concept prevalence for the influenza cohort. The columns are OMOP concept ID, count of patients’ visits with the concept, and prevalence of patients’ visits with the concept.

#### influenza_(non)hierarchical_concept_cooccurrence.txt

Hierarchical and non-hierarchical co-occurrence prevalence for the influenza cohort. The columns are the first OMOP concept ID, second OMOP concept ID, count of patients’ visits with the concept pair, and prevalence of patients’ visits with the pair of concepts.

#### influenza_symptom_prevalence.txt

Symptom prevalence for the influenza cohort. The columns are symptom name, the number of unique concepts having nonzero count in the symptom, count of patients’ visits with the symptom, and prevalence of patients’ visits with the symptom.

#### influenza_concept_prevalence_yearly_deviations.txt

The means and standard deviations of concept prevalence calculated based on the annual prevalence from the influenza cohort. The columns are OMOP concept ID, mean annual prevalence of the concept, and standard deviation of the annual prevalence of the concept.

#### influenza_concept_cooccurrence_yearly_deviations.txt

The means and standard deviations of concept co-occurrence prevalence calculated based on the annual prevalence from the influenza cohort. The columns are the first OMOP concept ID, second OMOP concept ID, mean annual co-occurrence prevalence of the concept pair, and standard deviation of the annual co-occurrence prevalence of the concept pair.

#### general_(non)hierarchical_concept_prevalence.txt

Hierarchical and non-hierarchical concept prevalence for the general cohort. The columns are OMOP concept ID, count of patients’ visits with the concept, and prevalence of patients’ visits with the concept.

#### general_(non)hierarchical_concept_cooccurrence.txt

Hierarchical and non-hierarchical co-occurrence prevalence for the general cohort. The columns are the first OMOP concept ID, second OMOP concept ID, count of patients’ visits with the concept pair, and prevalence of patients’ visits with the pair of concepts.

#### general_symptom_prevalence.txt

Symptom prevalence for the general cohort. The columns are symptom name, the number of unique concepts having nonzero count in the symptom, count of patients’ visits with the symptom, and prevalence of patients’ visits with the symptom.

#### general_concept_prevalence_deviations.txt

The means and standard deviations of concept prevalence calculated based on the annual prevalence from the general cohort data. The columns are OMOP concept ID, mean annual prevalence of the concept, and standard deviation of the annual prevalence of the concept.

#### general_concept_cooccurrence_deviations.txt

The means and standard deviations of concept co-occurrence calculated based on the annual prevalence from the general cohort data. The columns are the first OMOP concept ID, second OMOP concept ID, mean annual co-occurrence prevalence of the concept pair, and standard deviation of the annual co-occurrence prevalence of the concept pair.

#### concepts.txt

The definitions for all concepts used in the analyses. The columns are OMOP concept ID, concept name, domain, source vocabulary (the vocabulary that originally defined the concept, e.g., SNOMED, RxNorm, etc.), and concept class.

#### symptoms.txt

The definitions for all symptoms used in the symptom prevalence analysis. The columns are the symptom, OMOP concept ID, concept name, and domain.

### Technical Validation

#### Descriptive statistics

We analyzed CUIMC’s EHR data by calculating concept prevalence, concept prevalence ratio, concept co-occurrence, and symptom prevalence for three different cohorts, including the COVID-19 cohort. Summary statistics of the cohorts are provided in **Table 1**.

**Table 1.** Basic statistics of three cohorts in COHD-COVID. The statistics summarized here are based on the data from CUIMC as of September 1st, 2020.

|  | **COVID-19**<br>**cohort** | **Influenza**<br>**cohort** | **General**<br>**cohort** |
| --- | --- | --- | --- |
| Time range | 2020.03.01 –<br>2020.08.31 | 2014.01.01 –<br>2019.12.31 | 2014.01.01 –<br>2019.12.31 |
| # of patients | 4,127 | 3,261 | 175,930 |
| # of total inpatient visits | 4,846 | 3,860 | 314,680 |
| # of male patients<br>(# of inpatient visits) | 2,103<br>(2,518) | 1,454<br>(1,732) | 67,662<br>(128,908) |
| # of female patients<br>(# of inpatient visits) | 2,024<br>(2,328) | 1,807<br>(2,128) | 108,268<br>(185,772) |
| # of adult (18-64) patients<br>(# of inpatient visits) | 2,147<br>(2,511) | 1,315<br>(1,553) | 104,020<br>(173,843) |
| # of senior (65+) patients<br>(# of inpatient visits) | 1,980<br>(2,335) | 1,946<br>(2,307) | 71,910<br>(140,837) |

Concept prevalence and concept co-occurrence analysis on the COVID-19 cohort contain 1,066 concepts and 49,666 concept pairs, respectively, covering 290 condition, 705 drug, and 71 procedure concepts. Concept prevalence and concept co-occurrence analysis on the influenza cohort contain 1,392 concepts and 58,469 concept pairs, respectively, covering 510 condition, 756 drug, and 126 procedure concepts. Concept prevalence and concept co-occurrence analysis on the general cohort contain 11,250 concepts and 1,297,928 concept pairs, respectively, covering 4,362 condition, 3,428 drug, and 3,460 procedure concepts. Descriptive statistics of the results from the two analyses for each cohort are summarized in **Table 2** and **3**.

**Table 2.** Descriptive statistics of concept prevalence analysis for three cohorts.

| Domain | COVID-19 cohort |  | Influenza cohort |  | General cohort |  |
| --- | --- | --- | --- | --- | --- | --- |
|  | Total # of concepts | Mean (Min, Max) Prevalence | Total # of concepts | Mean (Min, Max) Prevalence | Total # of concepts | Mean (Min, Max) Prevalence |
| Condition | 290 | 0.0162<br>(0.0023, 0.3281) | 510 | 0.0200<br>(0.0028, 0.4990) | 4362 | 0.0018<br>(3.4956e-05, 0.2713) |
| Drug | 705 | 0.0285<br>(0.0023, 0.7654) | 756 | 0.0261<br>(0.0028, 0.5207) | 3428 | 0.0039<br>(3.4956e-05, 0.3554) |
| Procedure | 71 | 0.0452<br>(0.0023, 0.7947) | 126 | 0.0104<br>(0.0028, 0.0598) | 3460 | 0.0006<br>(3.4956e-05, 0.0427) |

#### Data quality analysis

Assessing data quality of EHR data is critical for its effects on secondary analysis for research in the healthcare and medical domains^14^. We calculated the sum of non-randomized counts of all concepts on a monthly basis for the COVID-19 cohort and on a yearly basis for the general and influenza cohorts. For each of the condition, drug, and procedure domains, we examined the total counts of concepts with the number of visits to detect any issues regarding data quality and temporal plausibility of the EHR data we used in the study^15^.

**Figure 2** shows the total counts across all concepts in the (**a**) condition domain, (**b**) drug domain, (**c**) procedure domain, and (**d**) the total visits per month between March 2020 and August 2020 for the COVID-19 cohort. The total counts of conditions, drugs, procedures and total visits all show steep increases in March and April 2020, when the number of COVID-19 cases surged in NYC, as shown in (**e**)^16^. The total counts of all domains and the total visits decreased starting May as the number of COVID-19 patients in NYC decreased.

**Figure 2.**
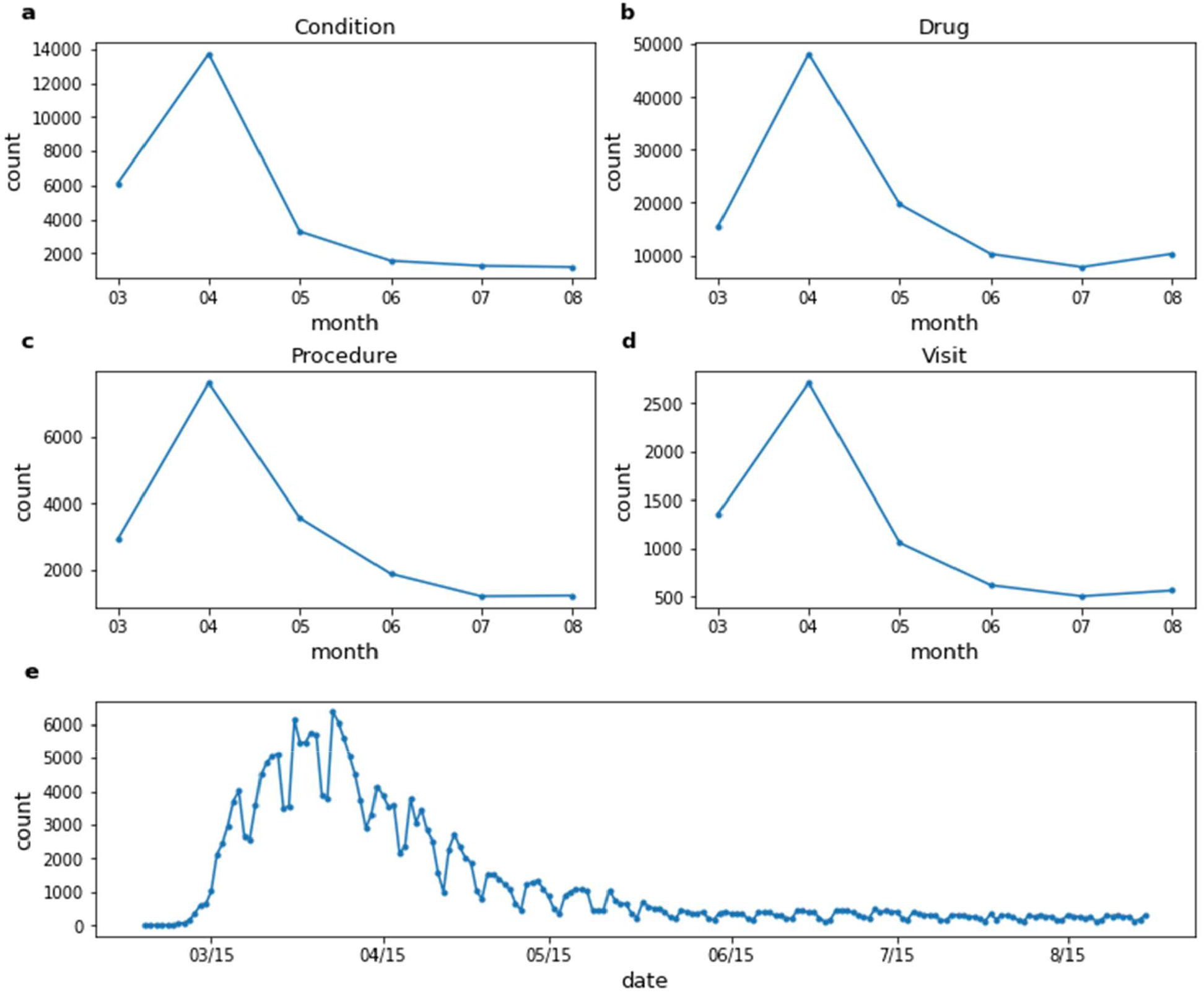
Total counts across all concepts in the (**a**) condition domain, (**b**) drug domain, (**c**) procedure domain, and (**d**) the total visits per month between March 2020 and August 2020 for the COVID-19 cohort. (**e**) Total COVID-19 positive cases per day in New York City from March 2020 to August 2020^16^.

A change in the EHR system can affect the quality and characteristics of EHR data collected for secondary research. CUIMC changed its EHR system as of February 1^st^, 2020 from Allscripts to Epic, which might affect the COVID-19 cohort data and its characteristics as compared against the influenza and general cohorts, which were collected prior to the EHR change. To detect the impact of the change, we performed a t-test for all concepts reported in the COVID-19 cohort between the counts from a pre-Epic period (January 1, 2020 – January 31, 2020) and from a post-Epic period (February 1, 2020 – Feb 29, 2020). The post-Epic period was chosen to minimize the inclusion of dates when COVID-19 would have impacted clinical practices in NYC. The counts of concepts were recorded on a daily basis. We considered there to be no difference between the two periods if the counts for the concept from the two periods are the same (i.e. all 0 counts for the two periods). Of all 1,066 unique concepts reported in the COVID-19 cohort, 119 (11.2%) concepts show a statistically meaningful difference (p-value ≤ 0.01) between the pre- and post-Epic periods. The p-values of the t-tests for all individual concepts are available in figshare^13^ to allow users to factor in these data quality considerations for each concept.

**Figure 3** shows the total counts per year across all concepts in the (**a**) condition domain, (**b**) drug domain, (**c**) procedure domain, and (**d**) the total visits per year between 2014 and 2019 for the general cohort and influenza cohort. The total counts of conditions, drugs, procedures and total visits per year for the two cohorts show consistent trends.

**Figure 3.**
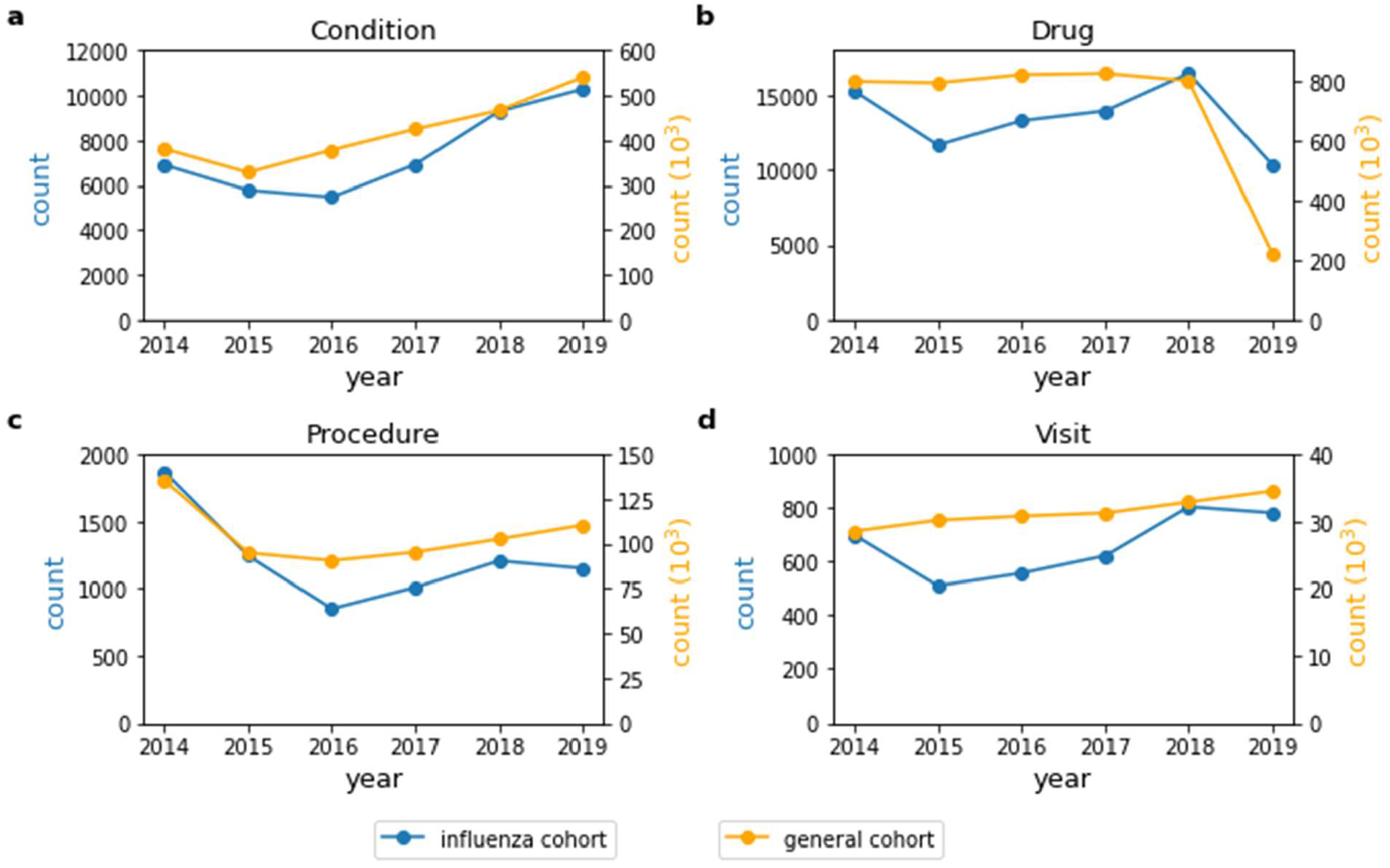
Total counts across all concepts in the (**a**) condition domain, (**b**) drug domain, (**c**) procedure domain, and (**d**) the total visits per year between 2014 and 2019 for the general and influenza cohort.

The annual mean and standard deviation of concept prevalence and concept co-occurrence for the general and influenza cohorts are available in figshare^13^ to assess the temporal variance of each concept and concept-pair. The mean and standard deviation of annual concept prevalence and co-occurrence should only be compared to each other to assess stability of the concept over the given time period of the dataset.

## USAGE NOTES

In this section, we show several motivating use cases to demonstrate the utility of COHD-COVID.

### Use case 1: Concept prevalence analysis on the three cohorts

Concept prevalence analysis on the three cohorts can be used to find clinical characteristics of the COVID-19 cohort. **Figure 4** shows the prevalence of 10 (**a**) condition and (**b**) drug concepts in the COVID-19 cohort, influenza cohort, and general cohort. We chose the 10 most prevalent condition and drug concepts in the COVID-19 cohort for this use case. For drug concepts, we chose the most prevalent drug ingredient concepts using hierarchical analysis to count multiple drugs having different brand names, dosages, and formulations but based on the same ingredients together. The condition concepts were chosen without hierarchical analysis to identify the top 10 specific conditions. Comparing the prevalence of these concepts between the COVID-19 cohort and the influenza or general cohorts provides contexual clues whether the concept is associated with COVID-19 or if its common among hospitalized patients. From **Figure 4** (**a**), we can confirm that *Fever, Cough, Dyspnea*, and *Acute lower respiratory syndrome* were more highly prevalent in the COVID-19 cohort than the two comparator cohorts. *Acetaminophen* was the most prevalent drug ingredient used for COVID-19 patients, which aligns with the high prevalence of *Fever*. We can see that *Hydroxychloriquine*, which was one of the drugs widely administered to COVID-19 patients^17^, showed high prevalence in the COVID-19 cohort.

**Figure 4.**
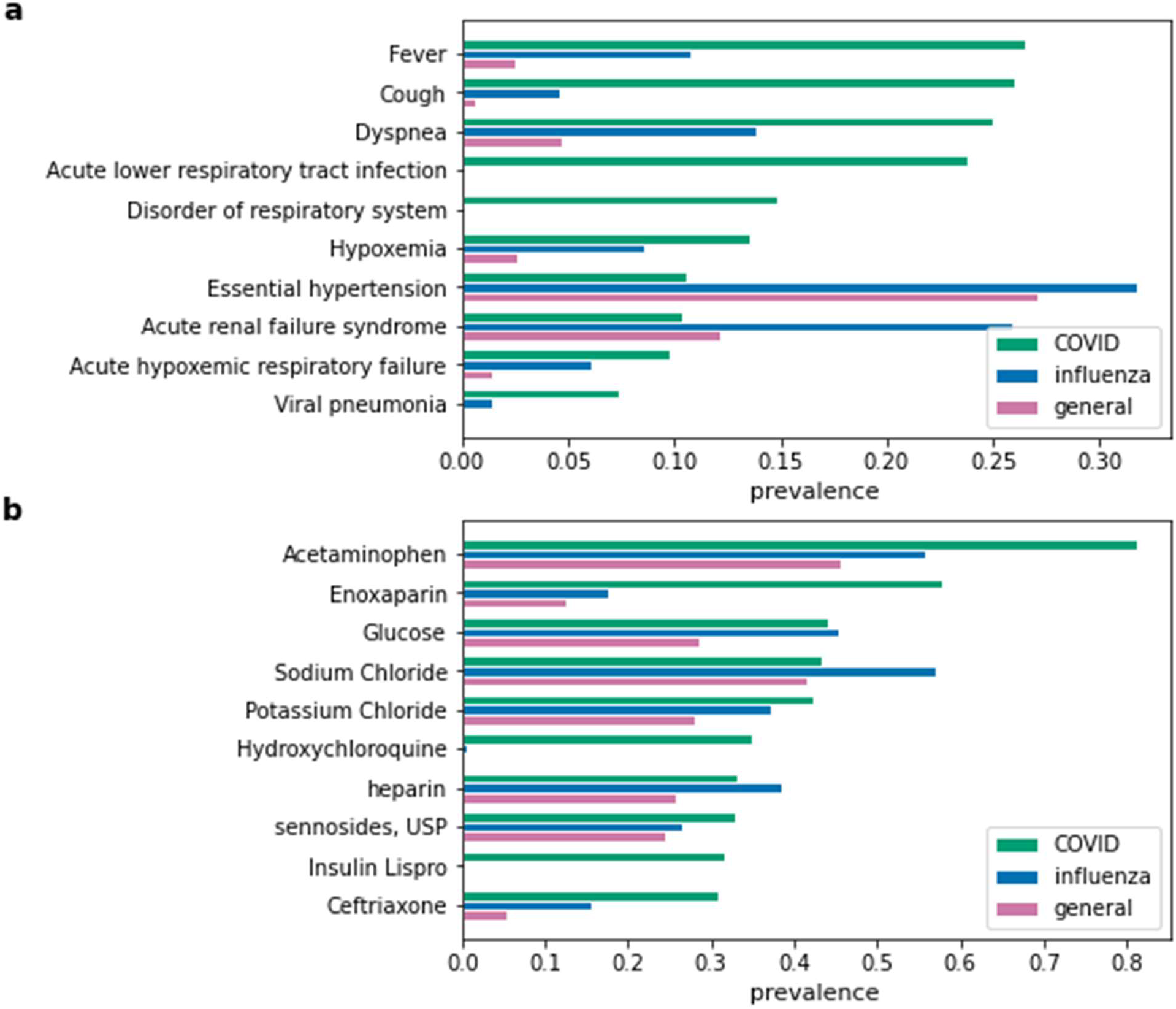
(**a**) Condition and (**b**) drug concept prevalence in the COVID-19 cohort, influenza cohort, and general cohort.

### Use case 2: Concept prevalence analysis on the COVID-19 cohort and its sub-cohorts

Concept prevalence analysis on the COVID-19 cohort and its sub-cohorts can be used to find clinical characteristics of the sub-cohorts. **Figures 5** and **6** show the prevalence of 10 (**a**) condition and (**b**) drug concepts, respectively, in the COVID-19 cohort stratified by gender and age. The 10 condition and drug concepts were the top 10 most prevalent concepts in the full COVID-19 cohort without stratification (the same concepts and prevalence calculation methods as in Use Case 1). **Figure 5** (**a**) shows that the senior cohort had higher prevalences in all 10 condition concepts than the full and adult cohorts. We can also confirm that *Essential hypertension* and *Hypoxemia* were particularly more prevalent in senior COVID-19 patients.

**Figure 5.**
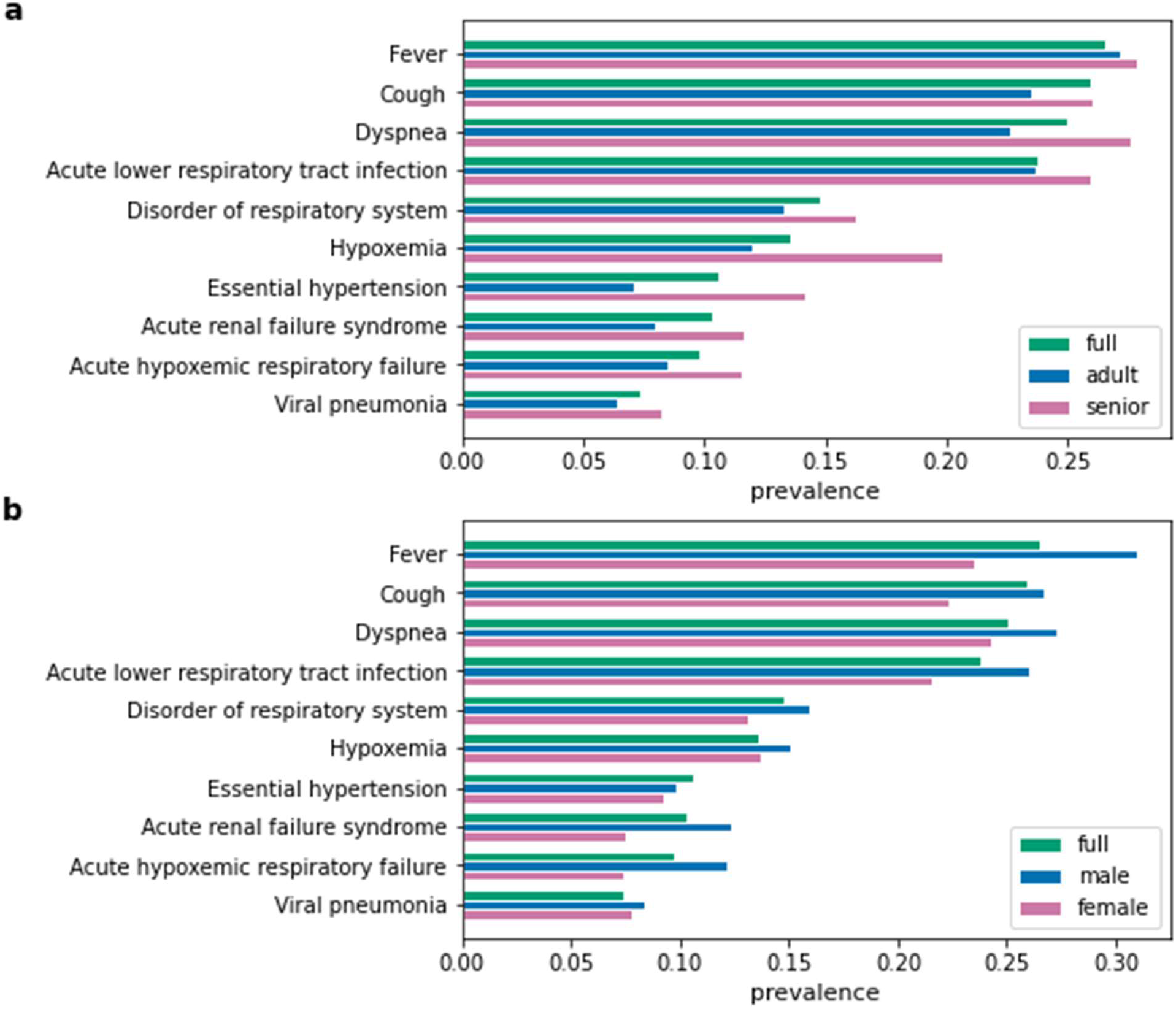
Condition concept prevalence in (**a**) age and (**b**) sex sub-cohorts of the COVID-19 cohort. The full COVID-19 cohort refers to the original COVID-19 cohort without stratification.

### Use case 3: Concept prevalence ratio analysis

Concept prevalence ratio analysis can be used to unveil how often specific concepts appeared in the COVID-19 cohort compared to the comparator cohorts. For example, **Table 4** shows the top 10 condition concepts that showed the highest concept prevalence ratio for the COVID-19 cohort relative to the general cohort and influenza cohort. High ratio of the concepts related to delivery was observed due to unusual spike of delivery events during the pandemic period from March 2020 to August 2020. It is interesting to see the high prevalence ratio of *Cerebral infarction* in the COVID-19 cohort compared to the influenza cohort, corroborating reports from several studies that severe acute respiratory syndrome coronavirus 2 (SARS-CoV-2) might be more likely to cause thrombotic vascular events, including stroke, than other coronavirus and seasonal infectious diseases^18,19^.

**Table 3.**
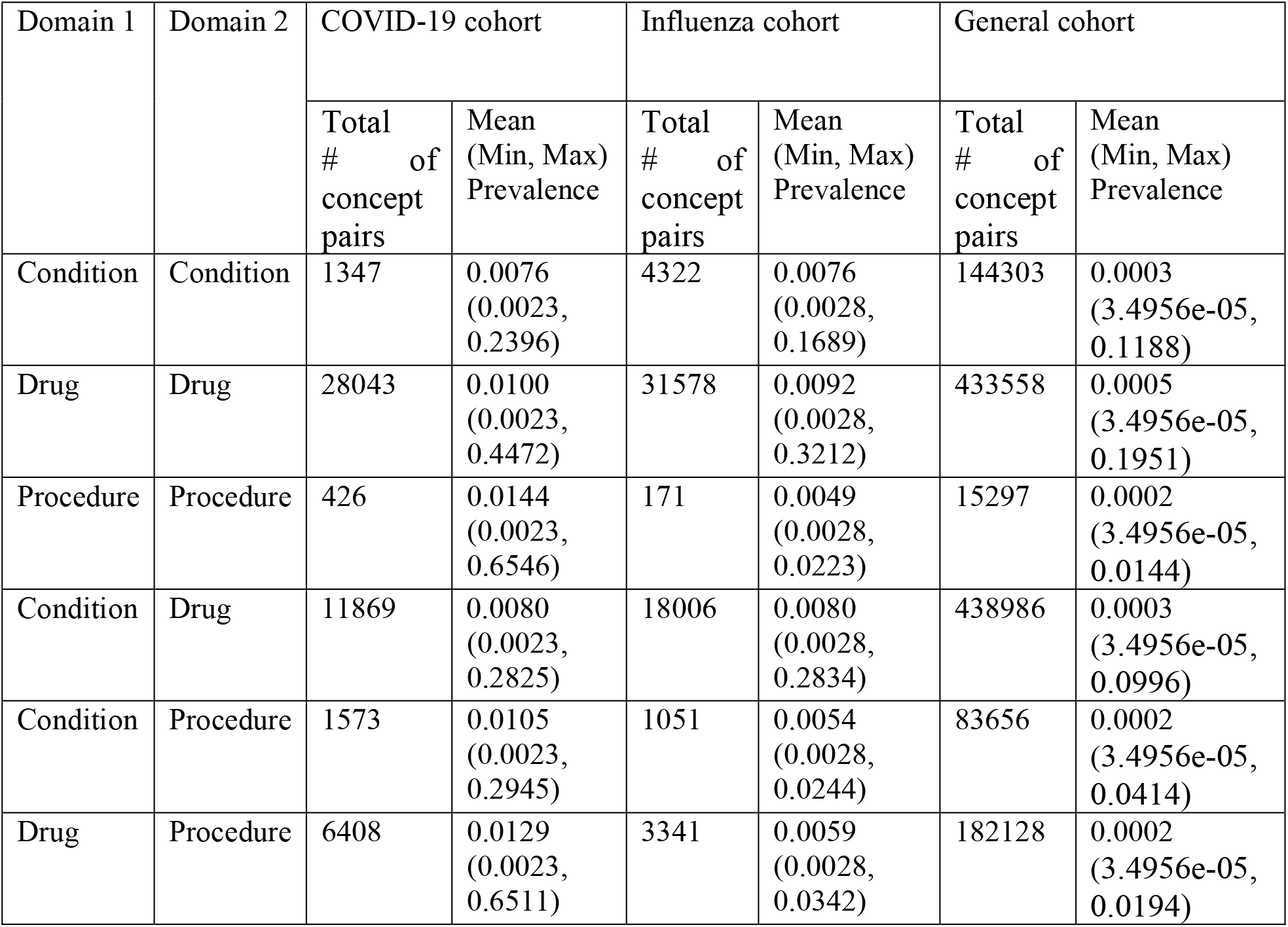
Descriptive statistics of concept co-occurrence analysis for three cohorts.

**Table 4.** Top 10 condition concepts that show the highest concept prevalence ratio for the COVID-19 cohort to the general and influenza cohort.

| Baseline cohort | Influenza cohort (prevalence ratio) | General cohort (prevalence ratio) |
| --- | --- | --- |
| 1 | Disease due to Coronavirus (3.09) | Outcome of delivery - finding (5.13) |
| 2 | Acute respiratory distress syndrome (2.04) | Chest pain on breathing (4.81) |
| 3 | Delivery normal (1.85) | Patient status finding (4.76) |
| 4 | Cough (1.74) | Disease due to Coronaviridae (4.75) |
| 5 | Acute respiratory distress (1.71) | Unplanned pregnancy (4.33) |
| 6 | Cerebral infarction (1.67) | Acute respiratory distress syndrome (4.01) |
| 7 | Disorientated (1.67) | Deliveries by cesarean (3.92) |
| 8 | Viral pneumonia (1.63) | Cough (3.84) |
| 9 | Hyperglycemia (1.38) | Acute lower respiratory tract infection (3.63) |
| 10 | Blood chemistry abnormal (1.34) | Acute respiratory distress (3.25) |

### Use case 4: Concept co-occurrence analysis of the COVID-19 cohort

Potential associations between specific concepts of interest can be found by co-occurrence analysis. For example, **Table 5** shows the top 10 most frequently co-occurring concepts with *Acute lower respiratory tract infection* in the COVID-19 cohort. We used non-hierarchical concept co-occurrence in this use case. The concepts associated with SARS-CoV-2 tests (i.e. *Disease due to Coronavirus, Radiologic examination, chest; single view*, and *Infectious agent detection by nucleic acid (DNA or RNA); severe acute respiratory syndrome coronavirus 2*) showed high co-occurrence prevalence ratio with *Acute lower respiratory tract infection. Hydroxychloroquine* also showed high co-occurrence prevalence ratio with *Acute lower respiratory tract infection*.

**Table 5.**
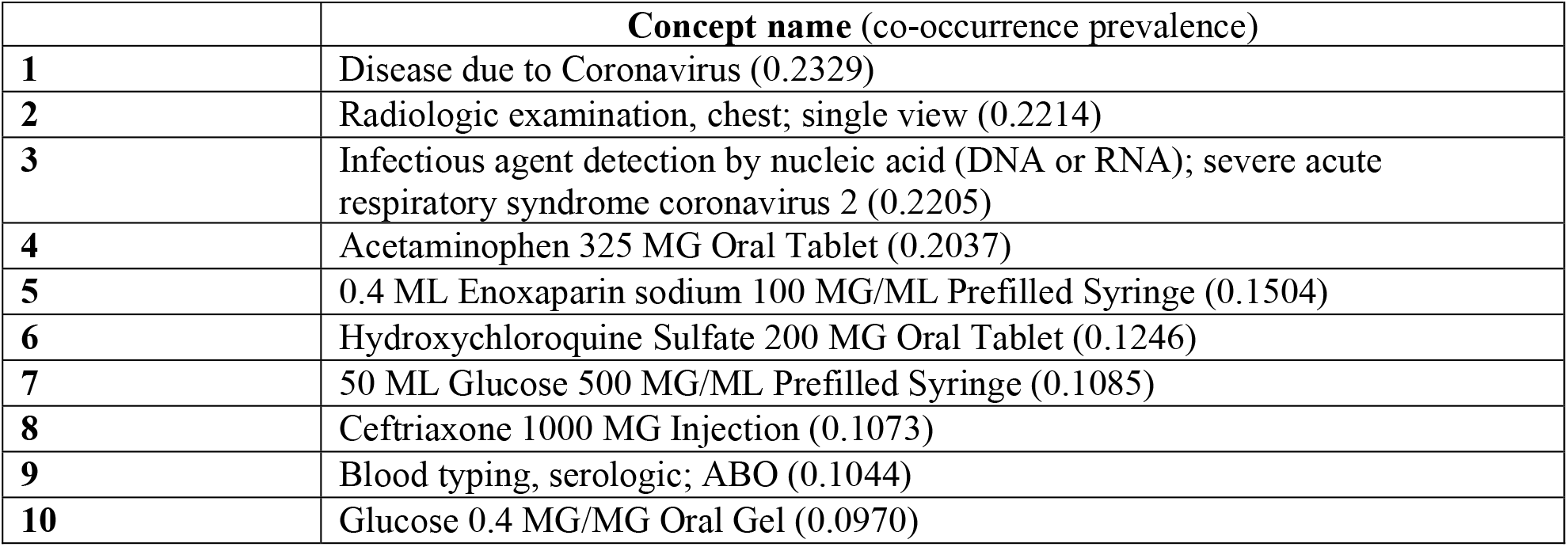
10 most frequently co-occurred concepts with *Acute lower respiratory tract infection* in the full COVID-19 cohort.

|  | **Concept name (co-occurrence prevalence)** |
| --- | --- |
| **1** | Disease due to Coronavirus (0.2329) |
| **2** | Radiologic examination, chest; single view (0.2214) |
| **3** | Infectious agent detection by nucleic acid (DNA or RNA); severe acute respiratory syndrome coronavirus 2 (0.2205) |
| **4** | Acetaminophen 325 MG Oral Tablet (0.2037) |
| **5** | 0.4 ML Enoxaparin sodium 100 MG/ML Prefilled Syringe (0.1504) |
| **6** | Hydroxychloroquine Sulfate 200 MG Oral Tablet (0.1246) |
| **7** | 50 ML Glucose 500 MG/ML Prefilled Syringe (0.1085) |
| **8** | Ceftriaxone 1000 MG Injection (0.1073) |
| **9** | Blood typing, serologic; ABO (0.1044) |
| **10** | Glucose 0.4 MG/MG Oral Gel (0.0970) |

### Use case 5: COVID-19 Symptom prevalence analysis

Symptom prevalence analysis can be used to examine symptom-level characteristics of the cohorts. **Figure 7** shows symptom prevalence of the 11 major COVID-19 symptoms for all three cohorts. The COVID-19 cohort showed higher prevalence in dyspnea, fever, and cough symptoms than the other two comparator cohorts, which aligns with the known characteristics and symptoms of COVID-19 patients^20^.

**Figure 6.**
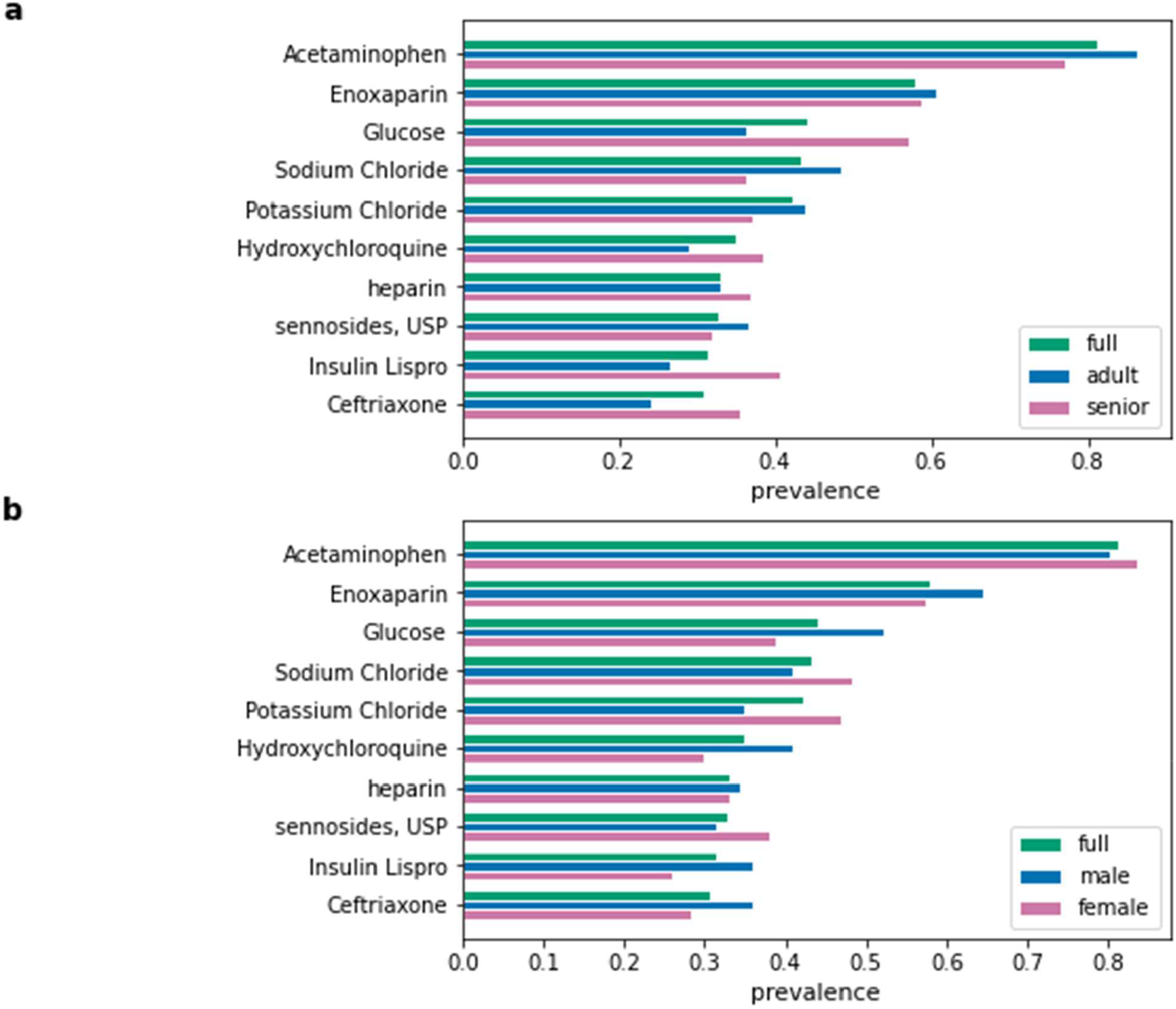
Drug concept prevalence in (**a**) age and (**b**) sex sub-cohorts of the COVID-19 cohort. The full COVID-19 cohort indicates original COVID-19 cohort without stratification.

**Figure 7.**
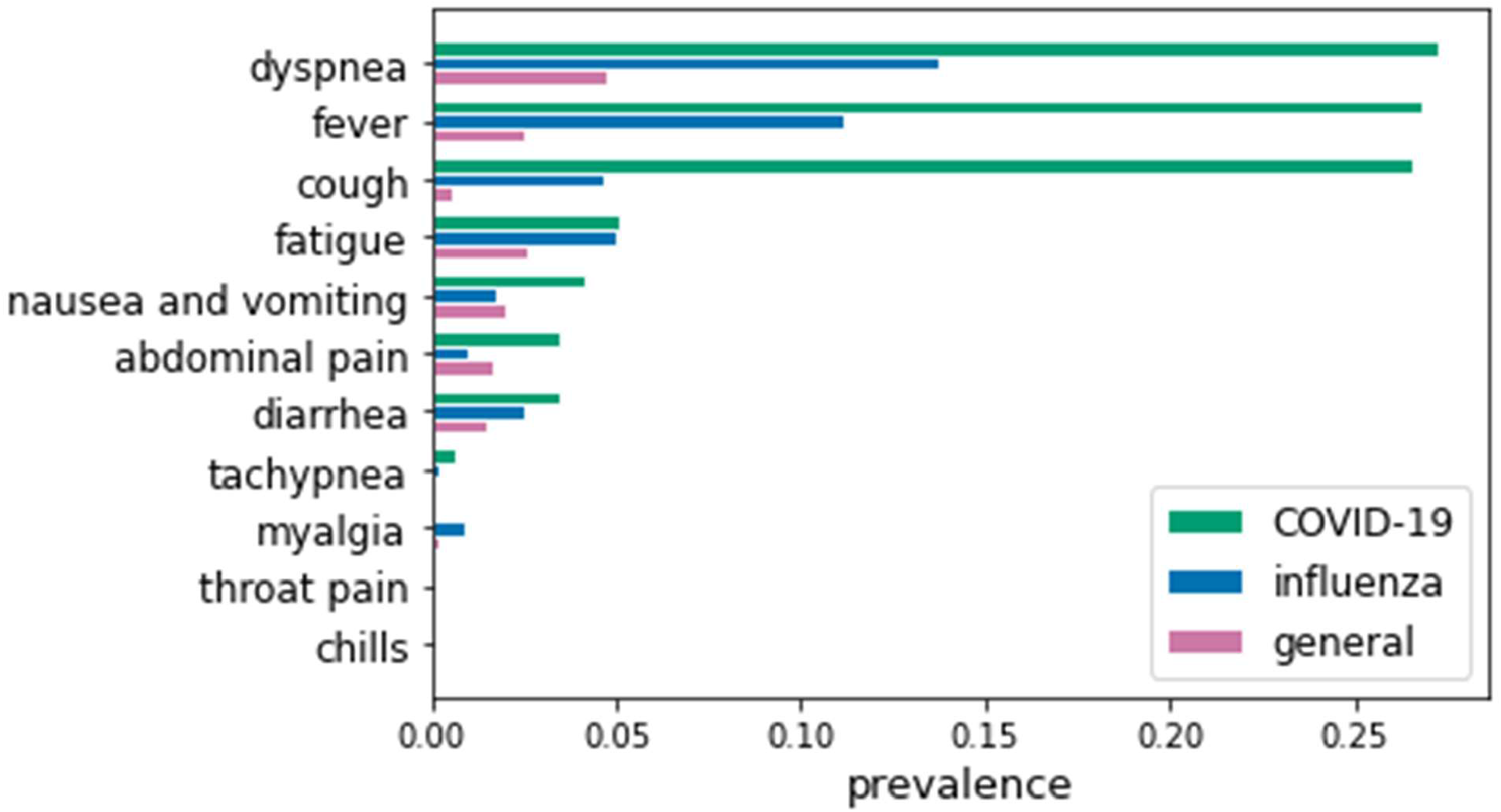
Symptom prevalence of 11 major symptoms in COVID-19 patients for all three cohorts.

## LIMITATIONS

There are several limitations to this work that should be noted. One of the limitations is that analyses performed in this work can be affected by the factors lying in the data acquisition process (e.g., change of the EHR system, and human biases and errors during entry). For example, coding trends and patterns (i.e., the trend and pattern of frequently used concepts) can be changed by a shift in system. Another limitation is that multiple visits from the same patient can be used to calculate the metrics in the analyses since we used visit-based count instead of patient-based count for the analyses. This could affect the results of the specific concepts appeared in the patients who made unusual visiting pattern.

The EHR data used in this study were obtained from a single site: the CUIMC campus of NewYork-Presbyterian (NYP). Even though NYP is the largest hospital in NYC covering the city and its surrounding area, we admit that performing the analyses based on the EHR data across multiple institutions and nations will be beneficial since multiple sites can diversify the population, improve accuracy, increase power and sensitivity to rare conditions, validate results by comparing across sites, and reduce variance that might exist in specific location. Since the OMOP CDM provides the fundamentals to perform the same analyses on clinical data across different sites, we hope to collaborate with future studies sharing clinical characteristics of COVID-19 patients and to generate a larger, richer, and more robust database that can be leveraged in translational research for COVID-19.

## Data Availability

The COHD-COVID API is available at http://covid.cohd.io/.

## CODE AVAILABILITY

The code to perform all analyses described in Methods and Usage Notes sections was written in Python 3.5.2 and is publicly available at https://github.com/Jayaos/cohd-covid. The COHD-COVID API is available at http://covid.cohd.io/.

## ACKNOWLEDGMENT

This work was supported by National Library of Medicine grant R01LM012895-03S1 and National Center for Advancing Translational Science grant 1OT2TR003434-01.

## AUTHOR CONTRIBUTIONS

Junghwan Lee and Casey Ta designed the study and implemented all methods and analyses. Jae Hyun Kim, Cong Liu, and George Hripcsak contributed to evaluate the methods and results of the analyses. Casey Ta and Chunhua Weng co-supervised the study. All authors were involved in developing the ideas and drafting the paper.

